# A Combined HLA Molecular Mismatch and Expression Analysis for Evaluating HLA-DPB1 *de novo* Donor-Specific Antibody Risk in Pediatric Solid Organ Transplantation: Implications for Solid Organ & Hematopoietic Stem Cell Transplantation

**DOI:** 10.1101/2020.01.06.20016709

**Authors:** Mengkai Shieh, Tristan J. Hayeck, Anh Dinh, Jamie L. Duke, Nilesh Chitnis, Timothy Mosbruger, Ryan P. Morlen, Deborah Ferriola, Carolina Kneib, Taishan Hu, Yanping Huang, Dimitri S. Monos

**Affiliations:** Department of Pathology and Laboratory Medicine, Children’s Hospital of Philadelphia, Philadelphia, Pennsylvania, United States; Department of Pathology and Laboratory Medicine, Perelman School of Medicine, University of Pennsylvania, Philadelphia, Pennsylvania, United States; Department of Pathology, Anatomy and Cell Biology, Thomas Jefferson University, Philadelphia, Pennsylvania, United States

## Abstract

**Background:** HLA molecular mismatch (MM) has been shown to be a risk factor for *de novo* donor-specific antibody (*dn*DSA) development in solid organ transplantation (SOT). HLA expression differences have also been associated with adverse outcomes in hematopoietic cell transplantation. We sought to study both MM and expression in assessing *dn*DSA risk.

**Methods:** One-hundred-and-three HLA-DP-mismatched SOT pairs were retrospectively analysed. MM was computed using amino acids (aa), eplets and, supplementarily, Grantham/Epstein scores. DPB1 alleles were classified as rs9277534-A (low-expression) or -G (high-expression)–linked. To determine the associations between risk factors and *dn*DSA, logistic regression, linkage disequilibrium (LD) and population-based analyses were performed.

**Results:** A high-risk AA:GX (recipient:donor) expression combination (X=A or G) demonstrated strong association with DP-*dn*DSA (p=0.001). MM was also associated with DP-dnDSA when evaluated by itself (eplet_p=0.007, aa_p=0.003, Grantham_p=0.005, Epstein_p=0.004). When attempting to determine the relative individual effects of the risk factors in multivariable analysis, only AA:GX expression status retained a strong association (RR=18.6, p=0.007 with eplet; RR=15.8, p=0.02 with aa), while MM was no longer significant (eplet_p=0.56, aa_p=0.51). Importantly, these risk factors are correlated, due to LD between the expression-tagging SNP and polymorphisms along HLA-DPB1.

**Conclusions:** The MM and expression risk factors each appear to be strong predictors of DP-*dn*DSA and to possess clinical utility; however, the two risk factors are closely correlated. These metrics may represent distinct ways of characterizing a common overlapping *dn*DSA risk profile, but they are not independent. Further, we demonstrate the importance and detailed implications of LD effects in risk assessment of *dn*DSA and possibly transplantation overall.

## 1. Introduction

The development of *de novo* donor-specific antibodies (*dn*DSA) after solid organ transplantation (SOT) is a major cause of significant organ loss (40%) within a 10-year period.^1–3^ Among multiple factors, the risk of developing *dn*DSA is related to human leukocyte antigen (HLA) mismatches between donors and recipients.^1,4,5^ In the context of organ shortage and the polymorphic nature of HLAs, absolute matching between recipients and donors is exceedingly difficult.

The definition of HLA mismatch in our field has evolved over time, based initially on serological (antigen) differences, then on incrementally more accurate assessments of HLA molecules by molecular techniques (evaluating antigen recognition domains), and finally, more recently, on epitope/amino acid differences (based on complete protein sequences). Relative degree of molecular mismatch (MM), whether expressed in amino acid or eplet units, has been shown to be an effective biomarker in assessing compatibility and predicting development of *dn*DSA.^4,5^

Another possible factor that may influence the development of *dn*DSA is the expression levels of mismatched epitopes in recipient and donor cells. Increased expression of HLA-mismatched alleles has been found to be associated with unfavorable transplantation outcomes.^6,7^ HLA-DP expression differences appear to influence the risk of graft versus host disease (GVHD) in HLA-DPB1 mismatched hematopoietic stem cell transplantation (HSCT).^7^ We hypothesized that an analogous but inverse phenomenon may be present in solid organ transplantation (SOT), in which HLA-DP-mismatched grafts with high-expression may elicit greater host immunogenic responses compared to low-expression grafts, particularly if the recipient has low expression DPB1 alleles that may render the immune system of the recipient less familiar with DP sequences and therefore more likely to perceive any DP sequence as non-self.

In order to investigate both the potential implications of differing grades of molecular mismatch as well as expression differences on development of *dn*DSA, the HLA-DPB1 locus was selected for study. HLA-DPB1 contains a molecular signature that allows assessment of expression level (low/high) for each allele. The 3’ untranslated region of the HLA-DPB1 gene contains a known single-nucleotide polymorphism (SNP), rs9277534 G/A, which is associated with either high (G) or low (A) expression of the gene in different cells and tissues.^7–10^

Utilizing well-established metrics for characterizing molecular mismatch, including amino acid, eplet, and physicochemical approaches^11^, we sought to ascertain the possible effects of both HLA-DPB1 molecular-structural mismatch as well as DPB1 expression on HLA-DP *dn*DSA development. While investigating these different associations, we observed an additional phenomenon, in which the inclusion of expression information (or more specifically, a SNP that tags expression) presents additional complexity. There is clear and expected overlap between the information captured by amino acid, eplet, and other measures of molecular mismatch; however, this particular rs9277534 SNP, which is used as a proxy for expression, appears to itself demonstrate non-random association with molecular mismatch. It is known in genome wide association studies that allele heterogeneity arising from multiple causal variants at a locus is frequently confounded by linkage disequilibrium (LD)^12^ or spurious correlation between alleles. In this analysis we found that both DPB1 molecular mismatch and expression appear to be informative metrics for predicting DP-*dn*DSA; however, there is non-random overlap in information captured between these two risk metrics. We show that the relationship between these two risk factors involves highly predictable patterns, based on LD and population allele frequency interdependencies. We also demonstrate that the relationships uncovered may additionally have important implications in the field of hematopoietic stem cell transplantation. Further investigation that takes into account the complexity of associations uncovered in this study is expected to contribute to unraveling and helping to identify foundational risk factors, their mechanisms, as well as their relative contributions to *de novo* DSA development and possibly GVHD as well.

## 2. Materials and Methods

### 2.1. Sample selection

Institutional review board (IRB) approval was granted by the IRB of the Childrens Hospital of Philadelphia (CHOP) for this retrospective study.

All solid organ transplants that took place between February 2013 and December 2019 and which were recorded in the HistoTrac database of the Children’s Hospital of Philadelphia were considered for this study. Exclusion criteria included: transplant pairs lacking two-field HLA-DPB1 typing, those involving null DPB1 alleles or DPB1 alleles with incomplete protein sequence in IMGT, and those matched at the DPB1 locus. Further excluded were patients who had pre-transplant HLA-DP DSA, multiple transplants recorded, or < 1 year between transplant and most recent DSA test. Following these exclusions, 103 transplant pairs remained for further analysis. HLA-DP DSA was considered positive if a donor HLA-DP Luminex single-antigen bead (or closest substitute) had a mean fluorescence intensity (MFI) ≥1500 at any time during post-transplant monitoring. Patients were monitored for DSA at regular intervals post-transplant (weeks to months initially, less frequently during the second year, and at least annually thereafter). Linkage information between HLA-DPB1 alleles and rs9277534 genotypes^13–15^ was utilized for classification into high-expression (rs9277534-G) and low-expression (rs9277534-A) DPB1 alleles. If a typing was ambiguous (a G-group designation), an approximation was used, analogous to that used for P-groups, as described in the section “Dependence between Molecular Mismatch and Expression Tagging Allele Combinations” below. The proportion of cases by transplanted organ was: 45/103 (44%) kidney, 43/103 (42%) heart, 14/103 (14%) lung, and 1/103 (1%) kidney-liver.

### 2.2. Assessment of Mismatches

An eplet is defined as a set of polymorphic HLA residues within a 3.0–3.5 Å radius on the molecular surface^16^ that theoretically constitutes the antibody binding site of the third complementarity-determining region of the immunoglobulin variable heavy chain.^17^ The eplets employed in this analysis were those contained in the HLA Epitope Registry v.2.0.^18^ Full-length amino acid sequences and alignments for HLA-DPB1 alleles were obtained from the IPD-IMGT/HLA Database v.3.38.0.^19^ A mismatch was counted when a donor amino acid or eplet had no matching counterpart on either of the patient alleles at the corresponding position. If both donor alleles possessed the same aa/eplet mismatch at the same position, the mismatch was only counted once.

Since there exist different ways to assess molecular mismatch, each with attendant advantages and disadvantages, this study analysed molecular mismatch based on two common approaches: simple enumeration of (1) eplet mismatches or (2) amino acid mismatches. We have further added a supplemental analysis that utilizes various physicochemical parameters to augment the basic molecular mismatch metric, as defined by the Grantham’s distance^20^ or Epstein coefficient of difference^21^ (Supplemental Methods and Results: Section 1, Figure S5, Supplemental Digital Content [SDC]). These measures are calculations based on amino acid properties that seek to provide a quantitative measure of the physicochemical change associated with specific amino acid substitutions in proteins. For both Grantham and Epstein metrics, the minimum score was used whenever a donor amino acid was mismatched to both patient amino acids at the same position, as a reflection of the degree of mismatch of that donor aa. Each individual score at the amino acid level was then added towards the final score at the transplant pair level.

### 2.3. Computational and Statistical Analysis

Our dataset was analyzed as a retrospective cohort study, with HLA-DP *dn*DSA development as the outcome of interest and a recipient:donor HLA-DPB1 expression–linked SNP genotype of AA:GX (homozygous AA low-expression recipient paired with a donor having at least one high expression G allele) or molecular mismatch as risk factors of interest. This AA:GX combination generally corresponds to the high risk DPB1 expression combination described by Petersdorf *et al*.^*7*^ in HSCT, in which they concluded that pairs with a low-expression A donor allele mismatched with a high-expression G recipient allele appear to be at elevated risk for GVHD.

A logistic regression model was fit to determine the marginal association of AA:GX (vs. non-AA:GX) status with development of HLA-DP *dn*DSA; then separately, the marginal association of molecular mismatch load (in terms of either amino acids or eplets) with DP-*dn*DSA development was tested with additional logistic regression models:

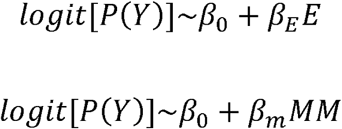

where the outcome *Y*_*i*_ is whether or not the recipient of transplant pair *i* developed DP-*dn*DSA, *E*_*i*_ is whether or not the transplant pair is of expression combination AA:GX, and *MM*_*i*_ is the molecular mismatch load (expressed in terms of amino acids or eplets).

Two additional multiple logistic regression models were fit, including AA:GX status and molecular mismatch (expressed in the first model as amino acid mismatch and in the second as eplet mismatch) as covariates to jointly assess their association with DP-*dn*DSA development:

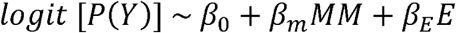

Since the 11% incidence of DP-*dn*DSA in our study population exceeded the 10% threshold^22^ that typically allows for the odds ratio (OR) to serve as a reasonable estimate of the relative risk (RR) in logistic regression analyses for cohort studies, we applied a correction—an approximation of the Mantel-Haenszel method^23,22^—which provides a better estimate of the relative risk when the outcome of interest is considered common (>10%).

DPB1 expression and molecular mismatch were chosen as the variables of focus in this study, without explicitly controlling for various other potentially relevant clinical covariates/risk factors, in order to avoid over-stratification of a modest dataset. However, both rs9277534 and molecular mismatch are biomarkers that are randomized from conception. Therefore, they allow for a closer approximation to a randomized trial, in which bias related to other covariates is reduced.^24,25^

All transplant pairs that passed our basic inclusion and exclusion criteria were included, which provides a relatively stable institutional treatment background from which the AA:GX and non-AA:GX pairs were selected. The AA:GX cohort consisted of 31 pairs, while the non-AA:GX cohort consisted of 72 pairs.

Transplant pairs were examined according to type of organ transplanted, with no clear aberrations in the general pattern observed (Figure S2, SDC). It was decided to evaluate all solid organ transplant data together, since stratifying by organ would result in relatively few cases for each organ type.

Computational procedures and data analysis were performed using custom programs written in Python v.3.7.4^26^ and R 3.6.1.^27^

### 2.4. Dependence between Molecular Mismatch and Expression Tagging Allele Combinations

To better understand the correlation between the patient-donor expression-SNP combinations and molecular mismatch load, the following procedure was performed: (1) Identification of the most frequent DPB1 alleles^28^ comprising the top 99% of the cumulative allele frequency, i.e. the common alleles, (2) linking of these common alleles to their respective rs9277534 genotypes by published linkage^13–15^ or based on exon 3 information^13^ (Tables S1, S2, SDC), (3) Permutation of every possible patient-donor DPB1 allele combination, (4) Calculation of corresponding molecular mismatch loads, and (5) weighting of results according to their associated population level allele frequencies. Since the reported allele frequencies were based on pre–April 2010 HLA nomenclature,^29^ the alleles were translated into the corresponding current P-groups and a representative allele was used for each P-group, which in most cases comprised >99% of the unambiguously typed alleles of the P-group, according to our institutional database of 11,132 high-resolution unambiguous DPB1 typings from all clinical and research databases (see Table S2, SDC, for details). For those P-groups with a less extreme split of constituent alleles, the alleles differed by a minimal amount of amino acids and the rs9277534 linkage was usually identical (Tables S1, S2, SDC). This procedure produced an analytical projection of the molecular mismatch distributions for every patient-donor rs9277534–SNP genotype. We termed this the frequency weighted permutation (FWP) dataset, in contrast to our Children’s Hospital of Philadelphia (CHOP) dataset. Mann-Whitney U tests (MW-U) were performed to assess the likelihood that the CHOP and FWP AA:GX vs. non-AA:GX samples were selected from populations with the same MM distribution.

The common alleles were further analyzed in a linkage disequilibrium (LD) analysis by extracting their sequences from the IPD-IMGT/HLA Database v.3.38.0.^19^ LD between each biallelic amino acid residue and the rs9277534 SNP was then computed^30^ (Table S3, SDC). Likewise, LD between the rs9277534 SNP and each HLA-DPB1 eplet defined in the HLA Epitope Registry v.2.0^18^ was computed by treating each eplet as a biallelic allele (either present or absent in each DPB1 sequence) (Table S4, SDC). The frequency of each polymorphic aa variant and eplet within each of the rs9277534-A vs.-G clades was also characterized, as shown in Figures S6, S7 (SDC).

### 2.5. HLA typing

HLA-DPB1 typing was performed as part of a general HLA typing procedure based on NGS technology (Holotype HLA, Omixon Biocomputing Ltd., Budapest, Hungary), as described previously.^31^

Despite the relevance of both DPB1 and DPA1 mismatches to DP-*dn*DSA development, DPA1 is known to be much less variable and polymorphic than DPB1 and not typically typed or used in transplant matching. As a result, it was not considered in the main study. We have, however, provided a supplemental analysis in which DPA1 is included, which decreases the number of available cases for analysis, but the general results appear similar to the main analysis (Supplemental Methods and Results: Section 2, SDC).

### 2.6. Accounting for SABs known to have a high false positive rate

Certain DP single-antigen beads have been described as having relatively high levels of false positives.^32^ The prevelance of DP alleles that correspond to the problematic beads was examined among the donors of DP-*dn*DSA positive and negative pairs. The prevalence of these problematic alleles was found to be no higher among the DP *dn*DSA+ pairs than among the DP *dn*DSA-pairs (Table S5, SDC).

## 3. Results

Within the set of 103 transplant pairs, 11 (10.7%) patients developed DP *dn*DSA, with a median follow-up of 34 months (range, 12–72). Figure 1 depicts molecular mismatch distributions for DP-*dn*DSA+ and DP-*dn*DSA− cases, with color-coding according to patient-donor rs9277534 A/G expression combinations. The AA-patient:GA-donor and AA-patient:GG-donor classes make up all but one of the DSA+ cases. These two groups collectively constitute a high-risk {low-expression recipient (AA) | high-expression donor (GX)} combination class, with AA representing homozygous low-expression and GX representing at least one rs9277534 high-expression (G)–linked allele. This high-risk patient-donor expression combination will hereafter be referred to as “AA:GX.” When fitting a simple logistic regression with DP-*dn*DSA development as the outcome of interest and AA:GX (versus non-AA:GX) as the risk factor of interest, a relative risk (RR) of 23.2 was observed (p-value=0.001) (Table 1).

**Table 1.**
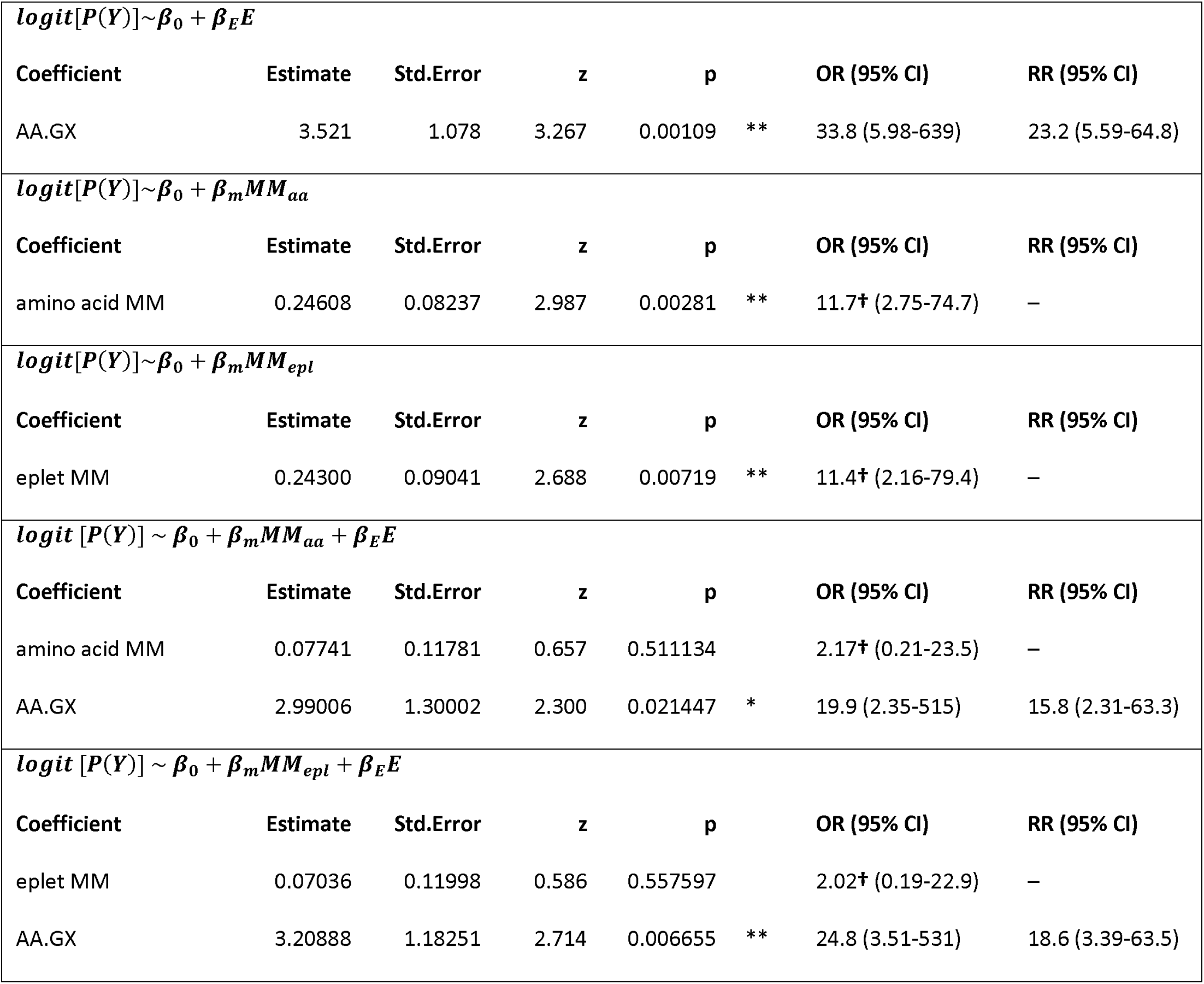
Testing association between molecular mismatch and rs9277534 with *dn*DSA. †The OR for molecular mismatch is calculated in terms of 10 mismatch increases. *p<0.05, **p<0.01. The RR estimation cannot be applied to the MM covariates in a straightforward and intuitive way, since it is meant to be applied to risk factors that involve a small number of discrete categories. Therefore, it has been omitted for the MM calculations, since the MM measures are represented in a ratio / count scale, each encompassing many values.

**Figure 1.**
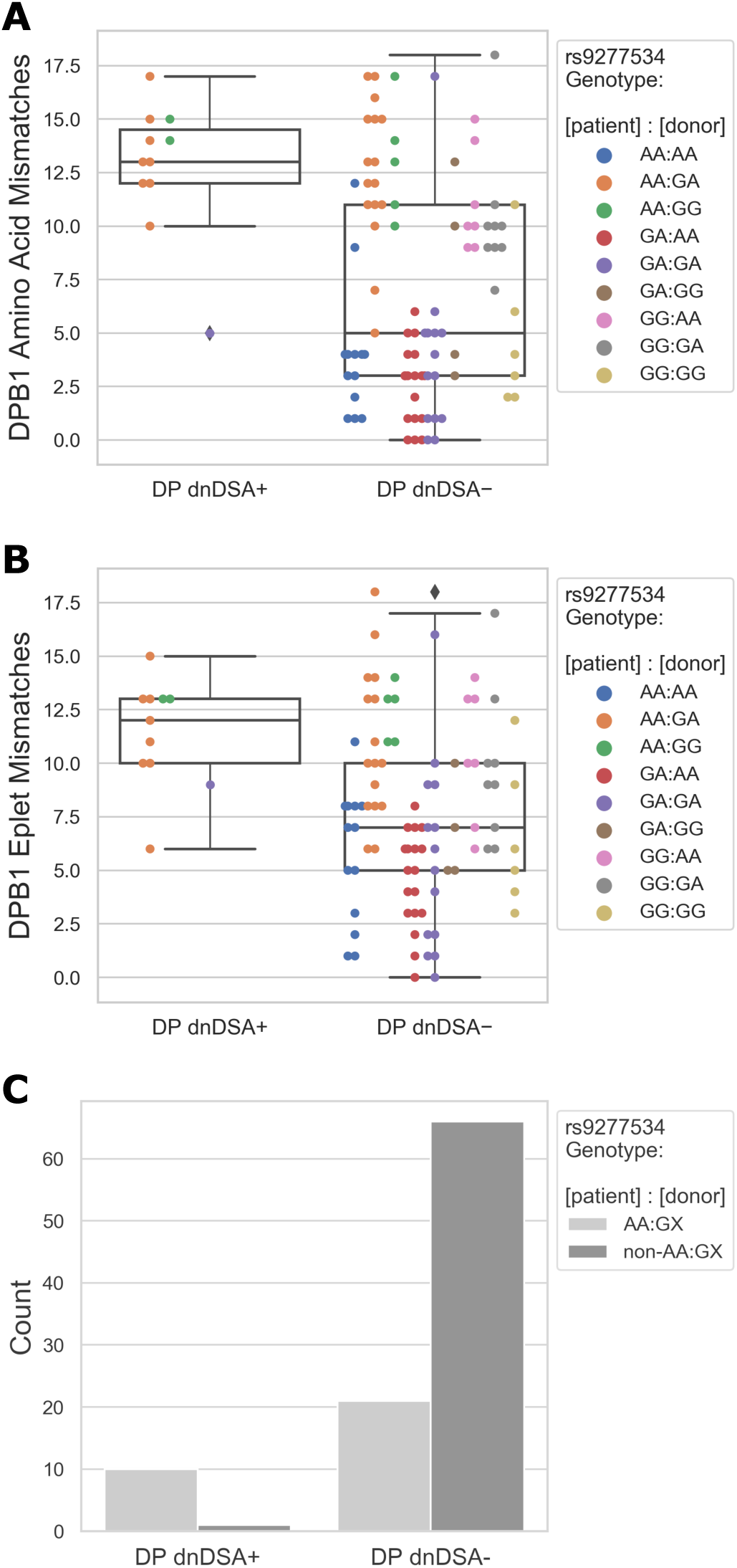
Distribution of molecular mismatch counts for DP dnDSA+/− groups, color-coded by patient:donor rs9277534 expression genotypes. (A) amino acid differences, (B) eplet differences. Results are color-coded by patient:donor rs9277534 A/G genotypes [A=low expression, G=high expression]. (C) Count of DP-*dn*DSA+ and DP-*dn*DSA− outcomes among AA:GX and non-AA:GX pairs.

The mean HLA-DPB1 amino acid mismatch for transplant pairs in DP-*dn*DSA+ and DP-*dn*DSA− categories was 12.7 and 7.1, respectively, while the mean eplet mismatch was 11.4 and 7.6, respectively. Both amino acid and eplet mismatch were associated with DP *dn*DSA development when evaluated using separate logistic regression models (p = 0.003 and 0.007, respectively; Table 1). It has been demonstrated that amino acid mismatch and eplet mismatch are correlated,^4^ which is also observed in our sample, with *r*^*2*^=0.81 (Figure S1, SDC). The differential distribution of molecular mismatch load for DP-*dn*DSA+ and DP-*dn*DSA− outcome groups is depicted in Figure 2.

**Figure 2.**
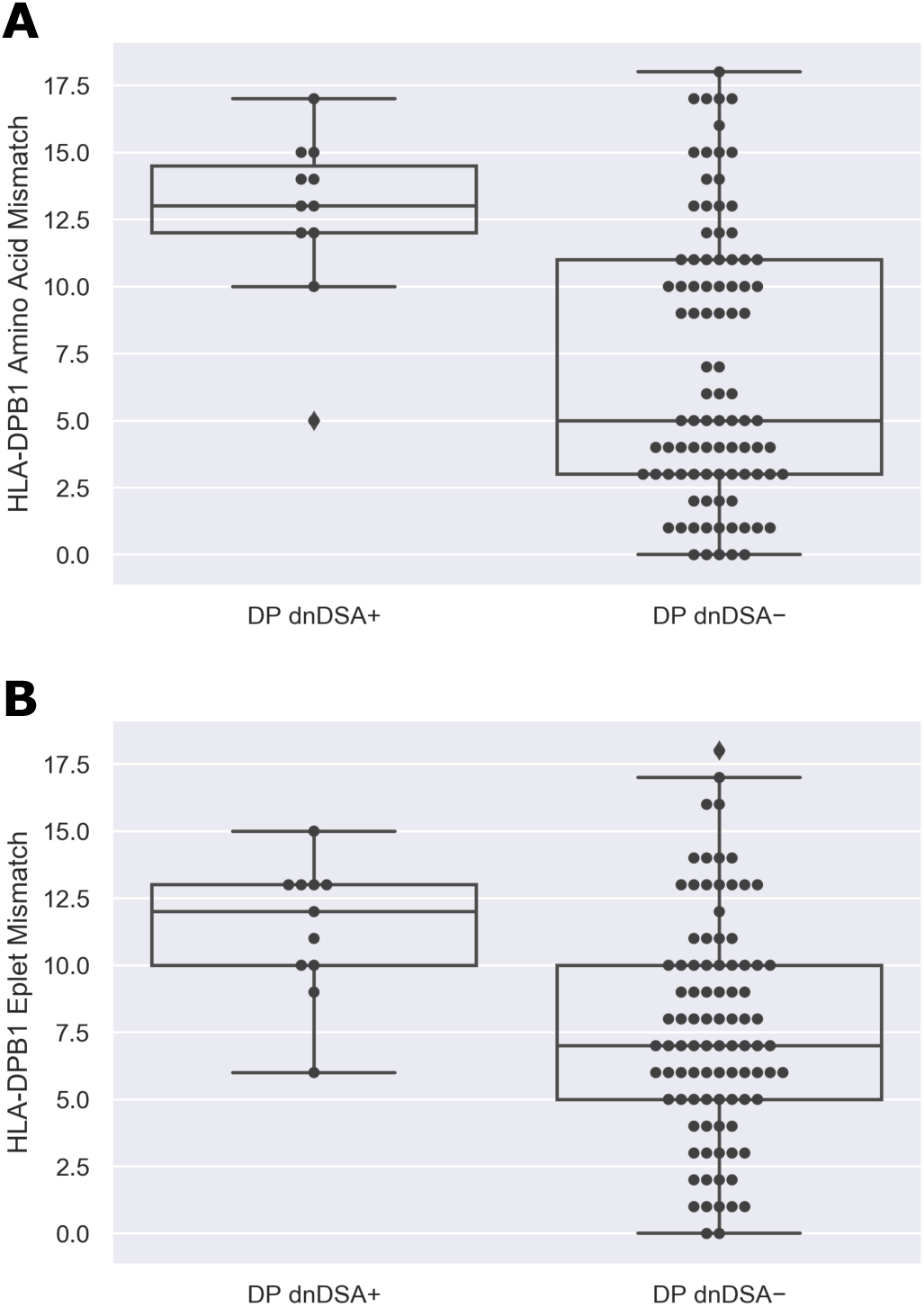
Molecular mismatch distributions according to DP-*dn*DSA+ and DP-*dn*DSA− outcome. (A) Amino acid and (B) eplet mismatch distributions according to DP-*dn*DSA+ and DP-*dn*DSA− outcome, expressed as box plots with individual points representing transplant pairs. The lower and upper box limits represent the first and third quartiles of the data, respectively, with the internal line indicating the median. The whiskers indicate the range of the data, out to a maximum of 1.5 times the interquartile-range beyond the upper and lower quartiles. Points more extreme than the whiskers are depicted as diamond-shaped outliers.

When combining patient-donor AA:GX status as a covariate together with either one of the molecular mismatch covariates in multiple logistic regression, we find that molecular mismatch is no longer associated with DP-*dn*DSA development (p=0.51 and 0.56 for the amino acid and eplet mismatch covariates, respectively), whereas AA:GX status is still significantly associated with DP-*dn*DSA development (p=0.02 or 0.007, when regressed with either the aa or eplet mismatch covariate, respectively). The relative risk of developing DP *dn*DSA+ for AA:GX pairs was estimated to be 15.8 (p=0.02) when analyzed together with the amino acid covariate and 18.6 (p=0.007) when analyzed together with the eplet mismatch covariate (Table 1).

When separately evaluating each of the two risk factors involving molecular mismatch and expression, both demonstrate significant association with DP-*dn*DSA. However, when we attempt to determine the individual and distinct effects of molecular mismatch vs. expression by computing their relative contributions at the same time (in multivariable regression), the effect of the AA:GX expression risk factor remains strong, but molecular mismatch is no longer significantly associated with DP-*dn*DSA. The instinct might be to interpret these results as indicating that expression is the more important factor. An important caveat in the above analysis is that the expression and molecular mismatch covariates are correlated, which is expected since the rs9277534 expression-tagging SNP and polymorphisms along the length of DPB1 are in linkage disequilibrium^13,33^ (Table S3, S4, SDC).

As a result of such a correlation, it is challenging to disentangle the true causal effects of expression and molecular mismatch. An interesting side effect of the LD, however, is that certain amino acid residues and eplets in high LD with the expression SNP (Table S3, S4, Figures S6, S7, SDC) represent nearly fixed amino acid/eplet differences between rs9277534 A and G allele types, resulting in a subset of specific and frequently occurring mismatched amino acids/eplets within the AA:GX pairs. These specific mismatches may potentially contribute significantly to the overall elevated DP-*dn*DSA risk among AA:GX pairs, and thus, their individual risk contributions will need to be disentangled also from the bare molecular mismatch enumeration and AA:GX classification risk factors themselves.

To assess the dependence between molecular mismatch and rs9277534-linked allele combinations, population level allele frequencies were leveraged (see Methods: Dependence between Molecular Mismatch and Expression Tagging Allele Combinations). An analytical projection of the molecular mismatch distributions for all the types of patient:donor rs9277534 genotype combinations was performed by permuting all possible patient:donor combinations of the 99% most frequent DPB1 alleles and weighting the results by the corresponding population level allele frequencies. The FWP molecular mismatch distributions were found to closely mirror the distributions of our CHOP-based dataset for every patient-donor rs9277534 expression combination (Figure 3, Table 2, Figure S3, SDC), which indicates that similarly constrained and characteristic molecular mismatch patterns would be expected to be observed for each patient-donor rs9277534–SNP genotype in any large study population. The distributions of MM across AA:GX category (AA:GX vs. non-AA:GX) within both CHOP and FWP datasets are significantly different (CHOP, AA:GX vs. non-AA:GX MW-U p<0.001; FWP, AA:GX vs. non-AA:GX MW-U p<0.001), whereas, when looking within AA:GX category but across datasets—CHOP vs. FWP—the distributions of MM do not appear statistically significantly different (AA:GX, CHOP vs. FWP MW-U p=0.29; non-AA:GX, CHOP vs. FWP MW-U p=0.31) (Figure 4).

**Table 2.** Mean ± SD of CHOP vs. FWP amino acid mismatch distributions for each Patient:Donor rs9277534 genotype.

| Patient:Donor rs9277534 genotype | CHOP aa MM (mean ± SD) | FWP aa MM (mean ± SD) |
| --- | --- | --- |
| AA:AA | 4.0 ± 3.2 | 3.4 ± 2.7 |
| AA:GA | 12.8 ± 3.0 | 11.8 ± 2.8 |
| AA:GG | 13.4 ± 2.4 | 14.3 ± 3.1 |
| GA:AA | 2.6 ± 1.8 | 2.7 ± 2.0 |
| GA:GA | 4.1 ± 4.1 | 5.2 ± 2.7 |
| GA:GG | 7.5 ± 4.8 | 6.3 ± 2.8 |
| GG:AA | 11.1 ± 2.4 | 10.4 ± 2.8 |
| GG:GA | 10.3 ± 3.1 | 11.6 ± 3.5 |
| GG:GG | 4.7 ± 3.4 | 6.9 ± 3.7 |
| non-AA:GX | 5.4 ± 4.3 | 5.8 ± 4.0 |
| AA:GX | 12.9 ± 2.8 | 12.4 ± 3.1 |

**Figure 3.**
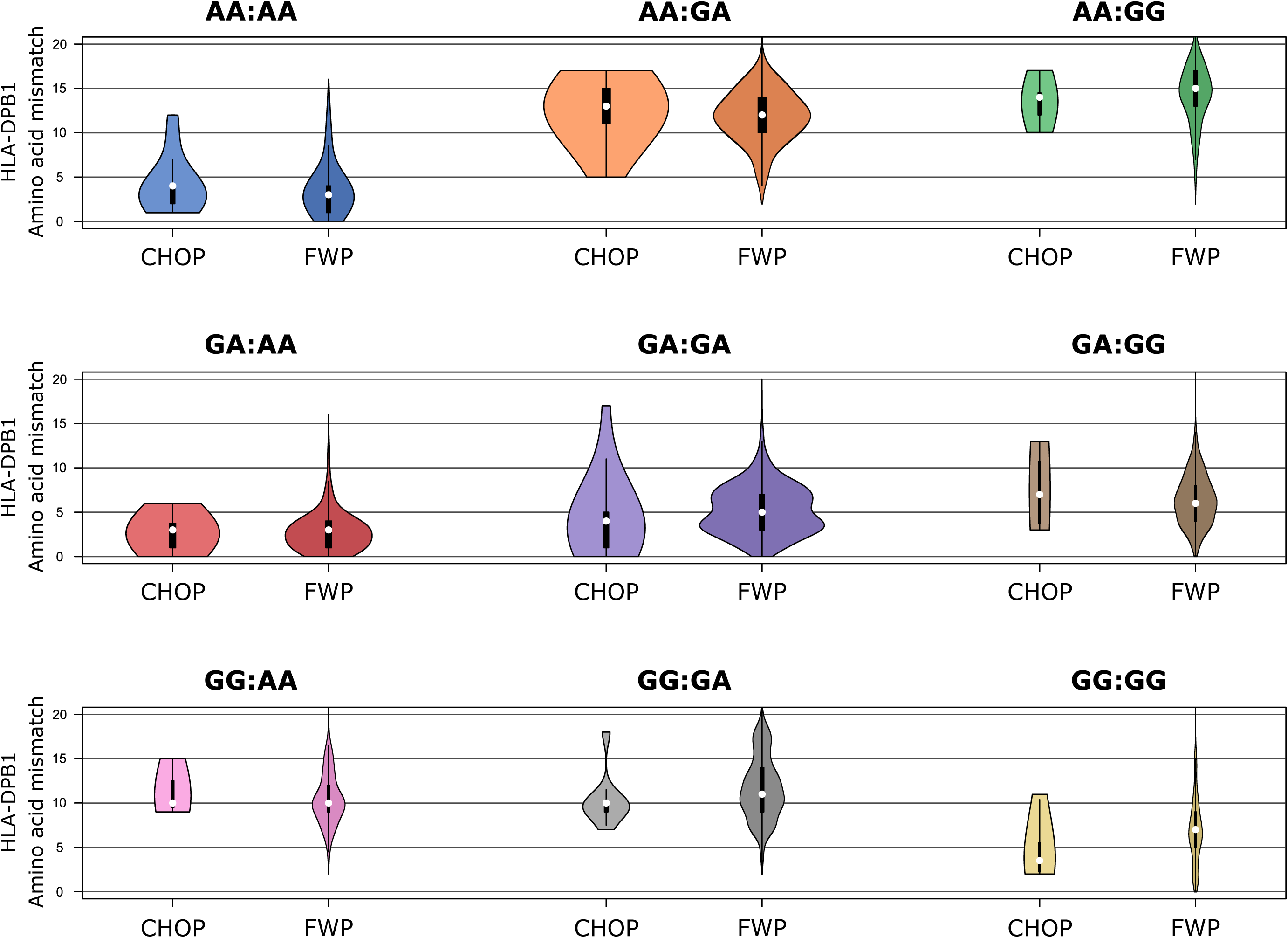
Frequency-Weighted Permutation (FWP) of most frequent patient-donor DPB1 alleles to define characteristic molecular mismatch distributions for each patient:donor rs9277534 genotype. An analytical projection of the molecular mismatch distributions for all the types of patient:donor rs9277534 genotype combinations was performed by permuting all possible patient:donor combinations of the 99% most frequent DPB1 alleles and weighting the results by the corresponding population level allele frequencies. The FWP molecular mismatch distributions are correlated with patient-donor rs9277534 expression genotypes in a way that closely mirrors the correlation seen in our CHOP population. Colors correspond to those of Figure 1 for ease of comparison. The width of each violin plot is proportional to the size of the data within it; these violin plots are therefore quantitatively comparable across the plots, within the same CHOP or FWP dataset. The lower and upper limits of the miniature boxes within the violin plots represent the first and third quartiles of the data, respectively, with the internal white dot indicating the median. The whiskers indicate the max/min of the data or 1.5 times the interquartile-range beyond the upper and lower quartiles, whichever is less extreme. The full range of the data is indicated by the upper and lower limits of the violin figures that enclose the box plots.

**Figure 4.**
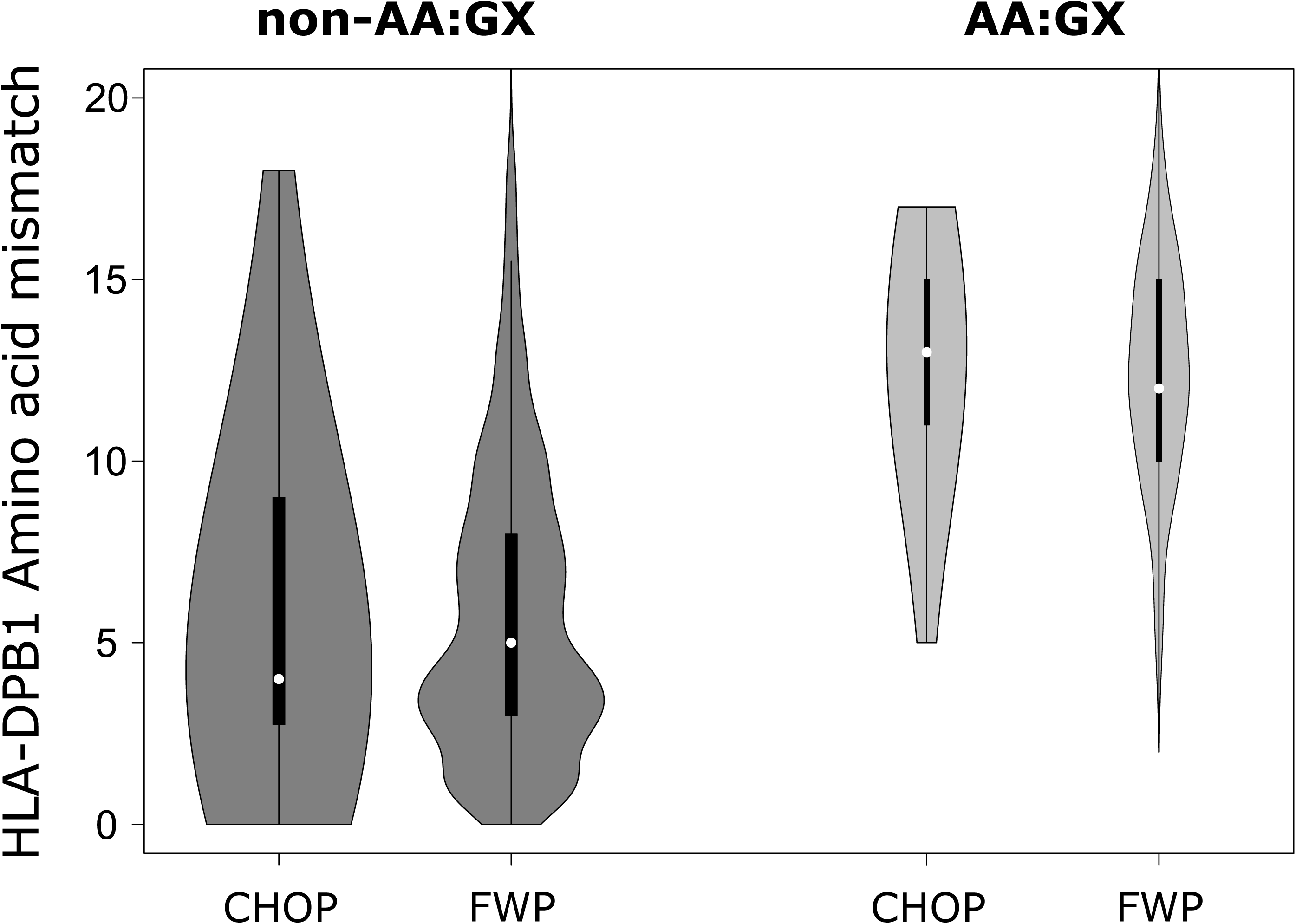
Frequency-Weighted Permutation (FWP) to define characteristic molecular mismatch distribution of AA:GX and non-AA:GX patient:donor genotype groups. The distributions of MM across AA:GX category (AA:GX vs. non-AA:GX) within both CHOP and FWP datasets are significantly different (CHOP, AA:GX vs non-AA:GX MW-U p<0.001; FWP, AA:GX vs non-AA:GX MW-U p<0.001); whereas, when looking within AA:GX category but across datasets—CHOP vs FWP—the distributions of MM do not appear statistically significantly different (AA:GX, CHOP vs FWP MW-U p=0.29; non-AA:GX, CHOP vs FWP MW-U p=0.31). The width of each violin plot is proportional to the size of the data within it; these violin plots are therefore quantitatively comparable across the plots, within the same CHOP or FWP dataset. The lower and upper limits of the miniature boxes within the violin plots represent the first and third quartiles of the data, respectively, with the internal white dot indicating the median. The whiskers indicate the max/min of the data or 1.5 times the interquartile-range beyond the upper and lower quartiles, whichever is less extreme. The full range of the data is indicated by the upper and lower limits of the violin figures that enclose the box plots.

To determine whether specific amino acid/eplet mismatches might clearly associate with the split of AA:GX cases into DP-*dn*DSA+ and DP-*dn*DSA− outcome groups, the specific amino acid/eplet mismatches were identified and compared between the AA:GX DP-*dn*DSA+ and DP-*dn*DSA− groups (Figure S4, SDC). There did not appear to be especially notable or divergent mismatch candidates identified.

To expand our molecular mismatch analysis to include methods that incorporate physicochemical parameters to attempt to quantify the degree of difference in each amino acid mismatch/substitution, we employed the Grantham^20^ and Epstein^21^ metrics for such a procedure. These methods appear to be potentially useful augmentations of the basic molecular mismatch metrics (Figure S5, Supplemental Methods and Results: Section 1, SDC).

## 4. Discussion

This work demonstrates that HLA expression analysis may have an important role to play in the assessment of immunological responses in SOT, beyond the traditional role of structural differences alone. However, disentangling the contribution of HLA expression from that of molecular-structural mismatch in *dn*DSA risk is not straightforward. Due to linkage disequilibrium, there are certain polymorphisms that are constrained to each of the rs9277534-A and -G expression-associated clades, which in turn constrain the possible molecular mismatch load of A/G patient-donor combinations. Even with larger sample sizes, in an attempt to better control for separate covariates, the problem of spurious correlation among molecular mismatch and expression is still expected to propagate, as is demonstrated through our analytical permutating of all common allele combinations (Figure 3). In such settings, machine learning techniques^34–36^ along with fine mapping^12,37^ and colocalization approaches^38,39^ specifically designed to address LD patterns and complex correlations could be adopted to hone in on particular causative mismatches and eQTLs. This would not only provide critical insight into potential true biologic influences that could be used to improve clinical testing but would also improve classification of benign mismatches, allowing for more flexibility in donor-patient matching.

The development of *dn*DSA remains a leading cause of chronic graft rejection/failure and a serious management challenge in SOT. Assessing the risk of *dn*DSA development can be instructive for pre-transplant assessment of donor-recipient compatibility as well as for post-transplant monitoring of DSA. The relevance and impact of the degree of HLA matching on *dn*DSA development, T-cell responses, and overall graft survival have been well described.^4,5^ Not much is known, however, as to whether the differential expression of different HLA loci and alleles within a particular locus play a role in graft survival and more specifically in either *dn*DSA development or T-cell immune responses. The realization that DP expression may affect the development of GVHD in HSCT suggests that a similar phenomenon may occur in SOT as well. In the setting of HSCT, HLA-DPB1 mismatch between a G-linked patient allele and an A-linked donor allele was shown to be associated with GVHD.^7^ In SOT, molecular mismatch has been established as a strong predictor of DSA development and subsequent graft failure.^4,5^ In this study, we do not attempt to dispute or affirm either of these concepts in the context of HLA-DPB1, but instead, we pose the interesting observation that the true causal factors of DP-*dn*DSA and their relative effect sizes from among DPB1 molecular mismatch and AA:GX expression are difficult to disentangle. When considered by themselves, both risk factors appear to be tagging *dn*DSA risk in our SOT cohort, but LD structure makes it difficult or impossible at the current sample sizes to differentiate true effects. It has been shown that the region from exon 3 to the 3’-UTR of HLA-DPB1, encompassing rs9277534, represents an evolutionarily conserved region and that stratified analysis of high and low risk expression groups and T-cell epitope permissive versus non-permissive mismatches is effective in risk assessment in the context of GVHD,^15^ but the possible interactions and underlying mechanisms have not been explicitly characterized. Further, it is possible that both the polymorphisms that underlie molecular mismatch and relative levels of expression are under coevolutionary selective pressures. These results should serve as motivation for future large-scale studies and development of enhanced techniques to better understand the individual effects and to better isolate true biologic factors.

We reflect on an example of the problem posed by this tangled association of DPB1 molecular mismatch and expression with transplantation-related outcomes. In the analysis of various DPB1-mismatched rs9277534–linked patient-donor allele combinations and their association with GVHD outcomes in HSCT, Petersdorf and colleagues^7^ conclude that “among recipients of transplants from donors with rs9277534A-linked HLA-DPB1, the risk of acute GVHD was higher for recipients with rs9277534G-linked HLA-DPB1 mismatches than for recipients with rs9277534A-linked HLA-DPB1 mismatches.” The comparison they are making is similar to comparing the Figure 3 orange AA:GA transplant pairs to the blue AA:AA pairs. If the AA:GA orange transplant pairs are reported to correspond to higher rates of adverse outcomes, an observer who lacks the molecular mismatch information may simply presume that such an outcome is due to expression-related effects. However, it is clear that the high molecular mismatch AA:GA orange distributions and the relatively low–molecular mismatch AA:AA blue distributions represent two quite opposite ends of the molecular mismatch spectrum. Therefore, there is good reason to believe that the Petersdorf *et al*. DPB1 GVHD study may also have been affected by such a tangled association between molecular mismatch and expression-tagging, given our FWP generated results. The FWP results should, by definition, generalize to any population with similar underlying DPB1 allele frequencies.

Quantity of amino acid mismatches in a sense undergirds both the direct T-cell epitope (TCE)^40^ and indirect PIRCHE^41^ methods of alloimmune risk stratification, since increasing the number of amino acid mismatches theoretically increases the chances of both an unfavorable TCE combination as well as PIRCHE score. Basic molecular mismatch may therefore correlate with both of these T-cell measures to some degree (we demonstrate such correlations using our FWP approach: Supplemental Methods and Results: Section 3, SDC). Both of these measures have been found to be associated with GVHD risk, each contributing in an independent capacity.^10,42,43^ Consequently, there is reason to believe that simple amino acid mismatch count may also have implications in HSCT and may correlate with adverse outcomes, as in SOT.

Our previous work involving computational assessment of miRNA targeting of the 3’UTR of HLA-DPB1 indicates that rs9277534 is likely only a marker of expression, in LD with causative factor(s) and not necessarily a causative SNP itself.^44^ There is a possibility that a number of SNPs in LD with rs9277534 may be driving the differential expression between the two major HLA-DPB1 clades^33^ through such mediators as miRNAs. Therefore, there may be additional layers of complexity to unravel when considering HLA expression effects, especially when dealing with other loci involving more complex expression-associated allele clades. There remains also a question of homozygous vs. heterozygous haplotype effects on expression and whether HLA haplotypes found in homozygous vs heterozygous individuals can display more complex interactions, such as those based on differentially encoded miRNAs per haplotype or various types of enhancer-promoter chromatin interactions. Besides the consideration of allele and haplotype-specific expression patterns of HLA molecules, tissue- and sex-specific expression differences can result in variable or even opposing expression patterns for one and the same SNP genotype.^45,46^ Therefore, a significant body of work remains as we attempt to develop a comprehensive understanding of HLA expression patterns and their role in HLA gene functions and histocompatibility.^47^

This study involved certain limitations. Firstly, it focused only on pediatric cases. Given that pediatric transplant cases are more limited in number and have population-specific factors that may influence *dn*DSA development,^48^ additional analysis of data from adult cases would be very appropriate. Additionally, DSA positivity in this study was based solely on a MFI threshold of 1500. As such, it is possible that the consideration of more subtle factors or criteria could have affected DSA assignment. Futher, this study did not control for certain clinical and patient variables that could contribute to bias such as sex, age, medication nonadherence, or physician-directed changes in immunosuppression (see Figure S15 though to see that sex is relatively balanced between DSA+/-groups but that controlling for all relevant donor-recipient sex/unknown combinations would likely have overstratified the dataset). It should be noted, however, that traditional sources of bias would not be thought to preferentially affect specific patient-donor DPB1 expression classes or molecular mismatch combinations in our study, as such molecular markers should be invisible to the clinician. Mendel’s law of segregation and independent assortment, often invoked in settings of Mendelian randomization, likely also apply in this case where the observed risk allele may be independent of other clinical covariates, offsetting the explicit need to control for such factors.^24,25^ One caveat, however, is that there are differences in frequencies of rs9277534 A and G alleles in different ethnic groups, which may contribute to bias. We would seek to control for such potential biases in future large-scale studies and see this as further motivation for more diverse study samples. A recent study by Philogene *et al*., however, suggests that such mixed-ethnicity factors may have less impact on *dn*DSA development than what might otherwise have been expected.^49^

As we work to disentangle and acquire a more nuanced understanding of additional and specific risk factors that may influence the development of *dn*DSAs or GVHD, the ultimate goal would be to integrate all relevant factors into a comprehensive risk analysis scheme. Specifically, this integrative approach would likely include both quantitative and qualitative components, reflecting numbers of epitope mismatches as well as immunogenicity of each specific epitope (whether based on physicochemical^11,20,50–53^ or other characteristics and/or capacity of epitopes to be presented by the responder’s HLA molecules^54,55^), levels of expression of these epitope-containing HLAs, as well as aspects of their regulation in various contexts. It is possible that additional metrics may be included in the future. This integrative approach should generate an improved system for assessing HLA mismatches and allow for a clearer determination of permissible mismatches and therefore influence both the longevity of transplants as well as the ability to perform a greater number of longer-lasting transplants.

## Data Availability

The data contained in this study can be made available upon reasonable request.

## ABBREVIATIONS

aa: amino acid
CHOP: Children’s Hospital of Philadelphia
*dn*DSA: *de novo* donor-specific antibody
FWP: frequency-weighted permutation
GVHD: graft versus host disease
HLA: human leukocyte antigen
HSCT: hematopoietic stem cell transplantation
IRB: Institutional Review Board
LD: linkage disequilibrium
MFI: mean fluorescence intensity
MM: molecular mismatch
MW-U: Mann-Whitney U test
NGS: next-generation sequencing
SAB: single-antigen bead
SDC: supplemental digital content
SNP: single-nucleotide polymorphism
SOT: solid-organ transplantation
TCE: T-cell epitope

## ACKNOWLEDGMENTS

We thank Steven D. Heron for contributions to quality control and critical comments. This work was possible thanks to the dedication of the histocompatibility technologists of the Immunogenetics Laboratory of the Children’s Hospital of Philadelphia.

## Notes

**3. Funding** This work was supported by institutional funds from The Children’s Hospital of Philadelphia to D.S.M.

### Competing Interest Statement

J.L.D., D.F., and D.S.M. receive royalties from Omixon Inc.

### Funding Statement

This work was supported by institutional funds from The Children’s Hospital of Philadelphia to D.S.M. Neither the authors nor their institutions received payment or services at any time from a third party for any aspect of the submitted work.

